# The antidepressant effects of vaporized N,N-Dimethyltryptamine: a preliminary report in treatment-resistant depression

**DOI:** 10.1101/2024.01.03.23300610

**Authors:** Marcelo Falchi-Carvalho, Handersson Barros, Raynara Bolcont, Sophie Laborde, Isabel Wießner, Sérgio Ruschi B. Silva, Daniel Montanini, David C. Barbosa, Ewerton Teixeira, Rodrigo Florence-Vilela, Raissa Almeida, Rosana K. A. de Macedo, Flávia Arichelle, Érica J. Pantrigo, Emerson Arcoverde, Nicole Galvão-Coelho, Draulio B. Araujo, Fernanda Palhano-Fontes

## Abstract

**Introduction:** N,N-Dimethyltryptamine (DMT), a naturally occurring psychedelic tryptamine contained in the indigenous ayahuasca brew has shown antidepressant effects. This Phase 2a clinical trial investigates for the first time the efficacy of isolated DMT in treatment-resistant depression (TRD).

**Methods:** Six TRD patients participated in an open-label, fixed-order, dose-escalation study, receiving a lower (15 mg) and then a higher (60 mg) dose of vaporized DMT in a single-day session. Depression severity was assessed using the Montgomery-Asberg Depression Rating Scale (MADRS) and the Patient Health Questionnaire-9 (PHQ-9) up to one-month post-dosing.

**Results:** Significant reductions in MADRS and PHQ-9 scores were noted from Day 1 to M1. The mean MADRS score variation from baseline to D7 was −22 points and −17 points at M1. PHQ-9 scores also showed significant decreases, mirroring the MADRS results. By D7, 83.33% of patients responded to treatment, with 66.67% achieving remission. At M1, 66.67% maintained response, and 50% maintained remission.

**Discussion:** The rapid onset and sustained antidepressant effects of vaporized DMT align with the paradigm of rapid-acting antidepressants to be used in the scope of interventional psychiatry. The non-invasive route and short-acting nature of DMT offer practical advantages, potentially enhancing accessibility to psychedelic treatments.

**Clinical Trial registration:** Clinicaltrials.gov NCT06094907

## Introduction

N,N-Dimethyltryptamine (DMT) is a naturally occurring psychedelic tryptamine, endogenous in small amounts in humans. When administered parenterally it induces profound changes in consciousness, including altered visual and auditory perceptions, frequently accompanied by deep emotional meaning, insights, significant shifts in thought processes, and mystical experiences, all condensed into an intensely powerful yet brief psychedelic experience^1,2^. Despite its potent psychedelic properties, the function and regulation of endogenous DMT within the human body is still a mystery. Although DMT is orally inactive due to the presence of monoamine oxidase (MAO) in the gut, South American Indigenous populations have long harnessed the psychoactive properties of DMT through Ayahuasca. This brew ingeniously incorporates monoamine oxidase inhibitors (MAOIs) to prevent DMT’s rapid deamination by MAO, thus enabling its psychotropic effects. In this case, the effects last a few hours, commonly including nausea and vomiting, besides the psychedelic effects^3^.

Recent clinical evidence has suggested the therapeutic effects of ayahuasca, particularly in the context of mood disorders^3,4^. The results from our Phase 2a and Phase 2b clinical trials with Ayahuasca in treatment-resistant depression (TRD) have been remarkable, suggesting rapid and sustained antidepressant effects in individuals with TRD^5–7^, a condition notoriously challenging to manage.

Following a comprehensive Phase 1 dose-escalation study, vaporized DMT proved safe and well-tolerated among healthy participants within the dosage range explored^8^. Encouraged by these findings, we have embarked on the first Phase 2a clinical trial to investigate the efficacy of pure DMT in treatment-resistant depression. By exploring this alternative route of administration, we aim to untangle the intricate relationship between DMT’s rapid psychedelic effects and its impact on depressive symptoms, contributing to the expanding horizon of psychedelic-assisted therapies.

## Methods

### Study design

This is an open-label, fixed-order, dose-escalation phase 2a clinical trial in patients with TRD. The study was approved by the Ethics Committee of the University Hospital Onofre Lopes (HUOL) (#45532421.0.0000.5292), registered at clinicaltrials.gov (NCT06094907).

### Participants

Candidates were recruited by physician referrals and were screened by a trained psychiatrist. Inclusion criteria: patients in a moderate/severe depressive episode according to MADRS (MADRS ≥ 20) during at least four weeks, and who have previously used at least two antidepressant medications, but without successful results (treatment-resistant, TRD). For exclusion criteria, see **Supplemental Methods**. Patients signed an informed consent before participation, maintained their pharmacological treatment, and abstained from psychedelics 14 days before dosing.

### Substance

DMT free base was vaporized using a device used for medical purposes in similar studies^8,9^ (for details, see **Supplemental Methods**).

### Procedures and Measurements

The study was conducted in the university hospital. After the screening, patients underwent psychological preparation. On dosing day (D0), they first received a lower DMT dose (15 mg), followed by a higher DMT dose (60 mg), 90 min after^1^. During the 15-minutes acute effects, patients remained lying down on a recliner with eyeshades and listened to music^2^. After the end of each dosing, patients underwent an immediate psychological integration, followed by questionnaires about the intensity, valence, and altered consciousness^10^ (ASC; see **Supplemental Measurements)**. Two hours after the second dosing, patients were evaluated by a psychiatrist and discharged with a family member or friend. One (D1) and seven (D7) days after the session, patients participated in follow-up integration sessions. At D0, D1, D7, D14, and one month (M1), depressive symptoms were evaluated by psychiatrists using the MADRS^11^ (Montgomery-Asberg Depression Rating Scale) and self-reports on the PHQ-9^12^ (Patient Health Questionnaire-9).

### Outcomes

The primary outcome measure was the mean change in depression severity assessed by the MADRS scale, comparing baseline with seven days after the dosing session. Secondary outcomes included the mean change in MADRS scores from baseline to D1, D14, and M1 after the dosing session; the response rate, defined as the proportion of patients meeting a reduction of 50% or more in baseline MADRS scores, and remission rates (MADRS⩽10); the mean change in PHQ-9 scores from baseline to D1, D7, D14, and M1 after the dosing session; changes in the ASC during the acute effects of DMT.

### Statistical analysis

Depression scores (D0 to M1) were analyzed in repeated measures General Linear Models (GLM; within-subjects factor = time; significance level = 0.05 (two-tailed)). *Post hoc* Dunnett’s multiple comparison test was used to compare each time point with the baseline values and effect sizes calculated by Hedge’s g. Analyses were performed with GraphPad Prism 7. The psychedelic experience measured by VAS and ASC scores (means (±SD) of dosing 2) are shown descriptively.

## Results

In this preliminary report, we included data from six patients who completed all assessments until M1. Patients’ sociodemographic, medication, and drug use are depicted in **Table 1**. For individual data, see **Table S1**. No participant showed critical physiological changes. Psychedelic experiences were subjectively intense (84.15±28.47), positive (30.40±19.02), and elicited mild effects in ASC total scores (35.17±14.38). Effects in each 5-ASC dimension are described in **Table S2**.

**Table 1.**
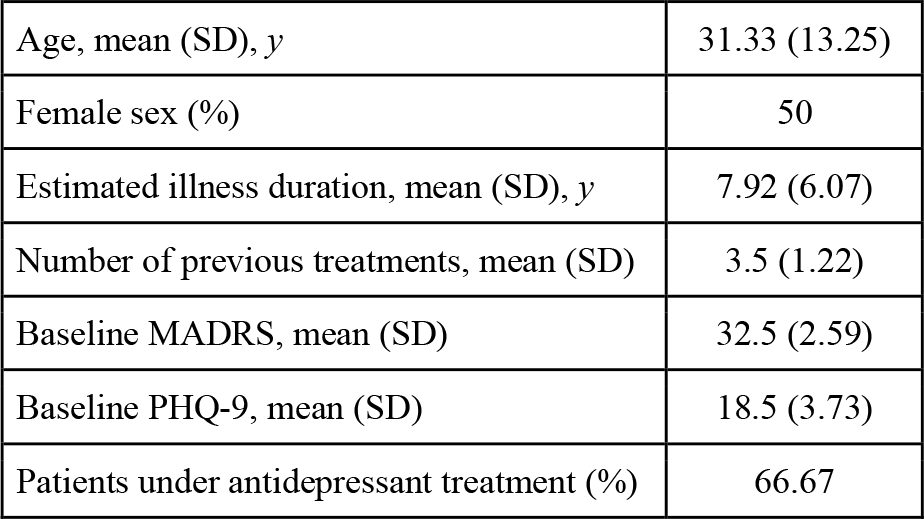
Demographics and clinical characteristics.

For MADRS scores, there was a main effect of time (*F*(4,20) = 10.41, *p* = 0.0001, *η*_p_^2^ = 0.67). Pairwise comparisons revealed a significant reduction of depressive symptoms at D1 (*p* < 0.0001; Hedge’s g = 4.70), D7 (*p*=0.0002; Hedge’s g = 2.89), D14 (*p*=0.0002; Hedge’s g = 3.34) and M1 (*p*=0.003; Hedge’s g = 1.80); **Figure 1A**). **Figure 1B** shows the proportion of patients who responded at D1: 5/6 (83.33%), at D7: 5/6 (83.33%), at D14: 5/6 (83.33%), and at M1: 4/6 (66,67%); and the remission rate at D1: 4/6 (66.67%), at D7: 4/6 (66.67%), at D14: 4/6 (66.67%) and at M1: 3/6 (50%). Individual % of change from baseline of MADRS scores at each time point are depicted in **Figure S1**.

**Figure 1.**
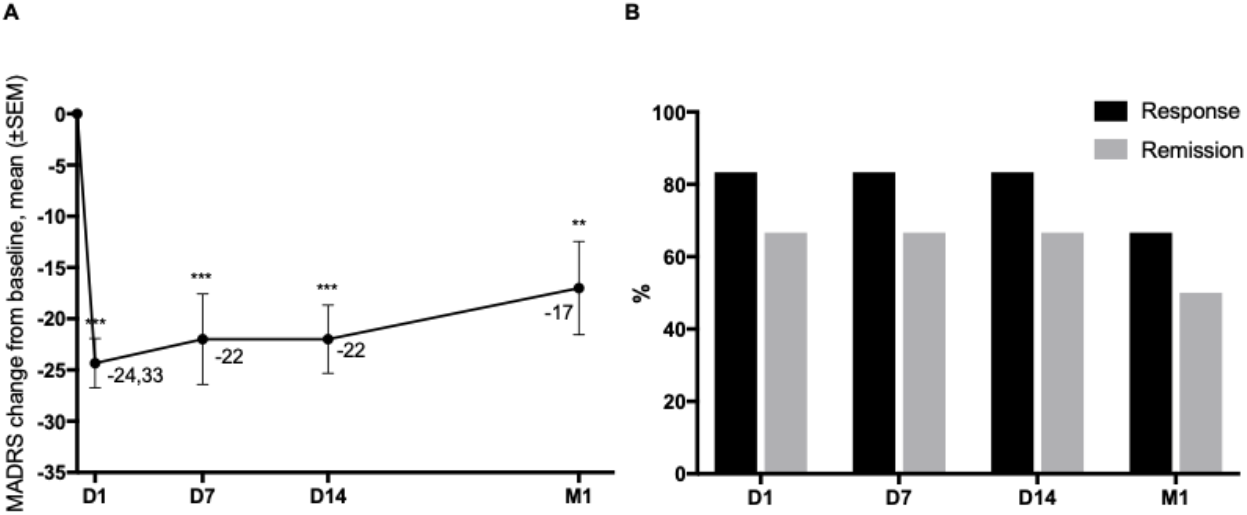
**A**) MADRS scores significantly decrease from one day, remaining significantly low until one month after the dosing session. **B**) Response and remission rates over time. ** p < 0.001; ***p < 0.0001.

For PHQ-9 scores, we also observed a main effect of time (*F*(4,20) = 6.27, *p* = 0.006, *η* _p_^2^ = 0.68). Pairwise comparisons revealed decreases, compared to D0, at D1 (*p* = 0.0008; Hedge’s g = 2.21), D7 (*p*=0.002; Hedge’s g = 1.69), D14 (*p*=0.003; Hedge’s g = 1.86) and M1 (*p*=0.014; Hedge’s g = 1.22); **Figure S2**.

## Discussion

This is the first clinical trial on the antidepressant effects of vaporized DMT in TRD. All patients showed improvement in their scores at different time points (D1, D7, D14, and M1) compared to the baseline. The rapid onset of antidepressant effects observed in our study, with significant reductions in MADRS and PHQ-9 scores as early as D1, aligns with the emerging paradigm of rapid-acting antidepressants.

No serious adverse events were observed during or after dosing. This is in line with what we observed in the phase I study^8^. During both sessions involving DMT, there was a rapid yet temporary increase in blood pressure and heart rate. DMT caused an acute rise in peripheral oxygen saturation. Overall, the findings suggest that our intervention with DMT is safe and tolerable for patients with depression.

A recent exploratory study on the antidepressant effects of intravenous DMT in seven patients with Major Depressive Disorder reported a moderate effect size (Hedge’s g = 0.75) with a mean reduction of 4.5 points in the Hamilton depression scale at D1^13^. This study lacks a specialized therapeutic environment and psychological support, factors that could have contributed to the modest outcomes regarding response and remission rates^13^. In contrast, our study achieved a higher effect size (Hedge’s g = 4.70) at D1, which was sustained for one month.

This disparity highlights the potential for enhanced therapeutic effects when psychedelic interventions are paired with psychological support in a thoughtfully designed setting. The superior response observed in our study may be attributed to this integrative approach, though variations in administration routes, dosages, and patient profiles are important considerations as well^14^.

Another recent study investigated the safety and efficacy of vaporized 5-methoxy-N,N-dimethyltryptamine (5-MeO-DMT) in sixteen patients with TRD^15^. They found a mean MADRS reduction at D7 vs. baseline of −24.4 points, whereas we found a mean reduction of −22 points (D7 vs. baseline). Both studies highlight the rapid antidepressant effects of short-acting psychedelic tryptamines using the same vaporizer device. However, our study differs in its use of DMT (vs. 5-MeO-DMT) and a single ascending fixed-dose approach, as opposed to an individualized dosing regimen. The positive outcomes in both studies reinforce the viability of using vaporized psychedelics as a promising therapeutic approach.

Furthermore, our data suggests the use of a short-acting psychedelic like DMT, as opposed to longer-lasting oral psychedelics (e.g., psilocybin, ayahuasca), maintains the therapeutic effects and offers practical advantages in terms of cost and operationalization, especially in settings requiring psychiatric and psychotherapeutic involvement. Along with the non-invasive inhalation route, contrasting to the intravenous methods, we believe lower the level of healthcare complexity required (secondary vs. tertiary or in-hospital care). Those aspects could be crucial in enhancing accessibility to psychedelic treatments and becoming suitable for public health systems, such as the Brazilian SUS (Unified Health System).

The open-label design and reduced number of patients in this study is a limitation, as it does not allow for the disentanglement of placebo effects from the results observed. Future directions include increasing the number of patients, assessing the need for repeated doses over time, and employing a more controlled experimental design (randomized and placebo-controlled). As we continue to understand the therapeutic potential of DMT, this preliminary study is poised to offer novel insights into the antidepressant capabilities of DMT, potentially transforming our understanding of its role in treating mood disorders.

## Data Availability

All data produced in the present study are available upon reasonable request to the authors.

## Author contributions

MFC, IW, NGC, DBA, and FPF designed the study; EA, DBA, and FPF acquired the authorizations; SRBS and EJP extracted and purified the substance; MFC and FPF selected the participants; MFC led the session, administered the substance and served as head of psychiatry for the study; HB, RB, and SL provided psychological support; DM, ET, and RFV served as psychiatrists to medical follow-up; RA, RKAM and FA provided nursing support; MFC, HB, RB, SL, DM, ET, RFV, RA, RKAM, and FA acquired the data; IW, NCG, DBA, and FPF provided research assistance; IW and FPF analyzed the data; MFC, IW, DBA, and FPF wrote the article; all authors reviewed the manuscript and approved the final version.

## Acknowledgments

We thank Raphael Egel for the production and composition of the music used in the study, Mariani Parra Cuerva for medical advice, Ayrton Senna Pinheiro for his support in chemical analysis, and João Arthur da Cruz Nunes, and Maria Luiza Assis for support in data pre-processing.

## Footnotes

1 The definition of these doses was based on our Phase I trial, which investigated 10 different doses across 5 schemes, ranging from 5 to 60mg^8^.

2 Produced and composed by Raphael Egel (instagram.com/liveenlightenment).

