## Supplemental for "The antidepressant effects of vaporized N,N-Dimethyltryptamine: a preliminary report in treatment-resistant depression"

**Supplemental Methods**

Participants

Exclusion criteria were: pregnancy (pregnancy test), no COVID-19 vaccination, heart, liver or kidney failure, asthma, coagulation diseases, uncontrolled hiper/hypothyroidism, severe obesity, uncontrolled systemic arterial hypertension, family suspicion or diagnosis of genetic monoamine oxidase deficiency, family history or diagnosis of cerebral aneurysm, history or diagnosis of heart rhythm disorders, valvular heart disease, chronic obstructive pulmonary disease (COPD), intracranial or cerebrospinal hypertension, epilepsy, or severe neurological disease, dissociative identity disorder, bipolar affective disorder, symptoms or family history of psychotic disorder, alcohol or drug use disorder, acute suicide risk, previous adverse response to psychedelic substances, flu-like symptoms, upper respiratory infection, condition that compromises understanding and performance of the measurements.

Substance

The DMT free base was vaporized in a vaporizer (Volcano® 'Medic 2', Storz & Bickel GmbH & Co, Tübingen, Germany) under 200°C for 10 sec in a 2-l balloon, following inhalation and retention for 3 sec. DMT purity (96.2%) was determined by gas chromatography-mass spectrometry (GC-MS) analysis.

Measurements

The psychedelic experience was measured on visual analogue scales (VAS) for intensity (0 = no effect, 100 = extremely intense effect), valence (-50 = extremely unpleasant effect; +50 = extremely pleasant effect; Wießner *et al*., 2021), altered state of consciousness (ASC; 94 items; 0 = No, not more than usual; 100 = Yes, much more than usual; Total score and 5 dimensions: as following: Oceanic Boundlessness (OBN); Anxious Ego Dissolution (AED); Visionary Restructuralization (VRN); Auditive Alteration (AA); Vigilance Reduction (VR)) (Dittrich, Lamparter, & Maurer, 2010).

**Supplemental Results**

Participants

Table S1. Demographic and clinical characteristics.

| Id | Age (range) | Sex | Estimated illness duration (y) | Past unsuccessful antidepressants (n) | Current antidepressants | Other psychotropic medication | Previous classic psychedelic substance use (times) | Baseline MADRS | Baseline PHQ-9 | Current psychotherapy |
|---|---|---|---|---|---|---|---|---|---|---|
| P1 | 36-40 | F | 1.5 | 2 | Venlafaxine 150mg | Zolpidem, 10 mg | 0 | 29 | 20 | no |
| P2 | 21-25 | M | 4 | 4 | Desvenlafaxine 50mg | none | 0 | 32 | 11 | yes |
| P3 | 21-25 | M | 9 | 2 | none | none | 0 | 36 | 20 | no |
| P4 | 21-25 | M | 11 | 5 | Desvenlafaxine 150mg | Aripiprazole 5 mg, Sodium valproate 1000 mg | 8 | 32 | 21 | yes |
| P5 | 26-30 | F | 4 | 4 | none | none | 0 | 31 | 20 | no |
| P6 | 51-55 | F | 18 | 4 | Escitalopram 10mg | Pregabalin 150 mg, Lithium 300 mg | 0 | 35 | 19 | no |

Table S2. Mean±SD scores of the five dimensions of the altered state of consciousness (ASC) after the second DMT dosing session.

| | |
|---|---|
| Oceanic Boundlessness (OBN) | 49.12±28.52 |
| Anxious Ego Dissolution (AED) | 12.13±7.98 |
| Visionary Restructuralization (VRN) | 55.43±20.03 |
| Auditive Alteration (AA) | 18.65±20.17 |
| Vigilance Reduction (VR) | 25.05±22.02 |

% change from MADRS baseline

| | D1 | D7 | D14 | M1 |
|---|---|---|---|---|
| P1 | 79,31 | 82,76 | 72,41 | 65,52 |
| P2 | 56,25 | 84,38 | 71,88 | 81,25 |
| P3 | 47,22 | 52,78 | 25,00 | 8,33 |
| P4 | 87,50 | 84,38 | 93,75 | 96,88 |
| P5 | 96,77 | 6,45 | 58,06 | 58,06 |
| P6 | 85,71 | 94,29 | 88,57 | 14,29 |

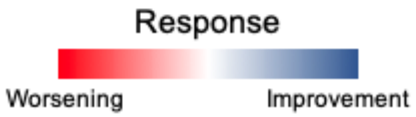


**Figure S1 -** Individual % of change from baseline of MADRS scores at each time point. All subjects improved at all time points. Positive responses appear in bluish, while negative would appear in reddish.

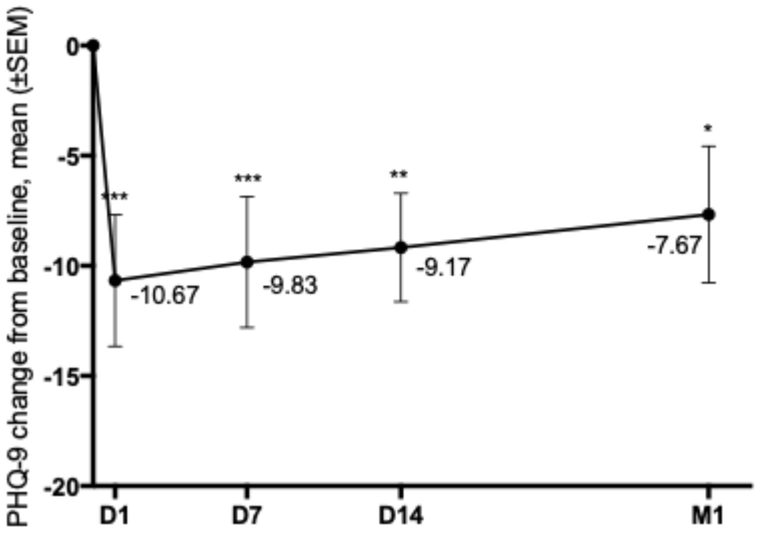


**Figure S2 -** PHQ-9 scores significantly decrease from one day, remaining significantly low until one month after the dosing session. $*p < 0.05$ $** p < 0.001$; $***p < 0.0001$.
